# MAGEC3 is a prognostic biomarker in ovarian and kidney cancers

**DOI:** 10.1101/2021.04.30.21256427

**Authors:** James Ellegate, Michalis Mastri, Emily Isenhart, John J. Krolewski, Gurkamal Chatta, Eric Kauffman, Melissa Moffitt, Kevin H. Eng

**Affiliations:** Department of Cancer Genetics and Genomics, Roswell Park Comprehensive Cancer Center, Buffalo, NY 14263 USA; Department of Urology, Roswell Park Comprehensive Cancer Center, Buffalo, NY 14263 USA; Department of Gynecologic Oncology, Roswell Park Comprehensive Cancer Center, Buffalo, NY 14263 USA; Department of Biostatistics and Bioinformatics, Roswell Park Comprehensive Cancer Center, Buffalo, NY 14263 USA

**Author notes:** **Corresponding author:** Kevin H. Eng, Ph.D., Department of Cancer Genetics and Genomics, Roswell Park Comprehensive Cancer Center, Elm and Carlton Streets, Buffalo, NY 14263 USA.

## Abstract

Rare variants in *MAGEC3*, members of the melanoma antigen gene family, are associated with BRCA-independent early onset ovarian cancers, while somatic mutations of this gene have been associated with kidney cancers. In this report, we quantified normal and tumor protein expression of MAGEC3 via immunohistochemistry in N=394 ovarian cancers and N=220 renal cell carcinomas. MAGEC3 protein levels fell into two categories – normal MAGEC3 and MAGEC3 loss – characterized by expression equivalent to normal tissue or significantly lower than normal tissue, respectively. Interestingly, cases with MAGEC3 loss demonstrated better overall survival in both ovarian cancers and renal cell carcinomas, which resembles patient outcomes with BRCA2 loss. MAGEC3 protein expression was associated with upregulation of pathways regulating G2/M checkpoint (NES: 4.13, FDR<0.001) and mitotic spindle formation (NES: 2.84, FDR<0.001). Increased CD8+ cell infiltration, coordinate expression of other cancer testis antigens, and tumor mutational burden were also associated with MAGEC3 expression. To emphasize the impact of these results, we built a prognostic RNA-based model using N=180 cancers of an independent cohort with matching transcriptomic data and tested its performance in two large public cohorts (N=282 ovary and N=606 kidney). Results based on predicted protein scores within these patients validated those discovered in patients with directly measured MAGEC3 protein. The RNA model was reproduced in independent cohorts implying a broader potential for MAGEC3-driven disease etiology and relevance to potential treatment selection.

**STATEMENT OF TRANSLATIONAL RELEVANCE:** MAGEC3 protein is expressed in multiple tissues and is dysregulated in cancer. In this work, we show that ovarian and kidney cancer patients with loss of MAGEC3 protein have favorable prognosis, indicating that MAGEC3 protein level may be used as a prognostic biomarker. Integrative genomic analysis of patients spanning more than nine cancer types showed an association between MAGEC3 protein and genes affecting stress response, including those involved in cell cycle and DNA damage repair. Additionally, it is correlated with tumor mutational burden in patients with mutated oncogenes. These associations suggest that MAGEC3 protein levels may be used to identify patients with deficient DNA damage repair mechanisms that can be targeted by PARP inhibitors. To operationalize this idea, we use machine learning to predict MAGEC3 protein levels from RNA sequencing data which can facilitate the identification of patients for treatment stratification according to their MAGEC3 status.

## INTRODUCTION

We previously reported the linkage between early onset, BRCA-independent ovarian cancers and a locus on Xq27.2 that contains the gene *MAGEC3* [1] identified from inheritance patterns in the Familial Ovarian Cancer Registry (Buffalo, NY) [2] that heavily favored clustering among sisters whose father transmits the risk allele [3]. A separate report leveraged sex-imbalanced mutations in *MAGEC3* sporadic kidney cancers [4] to argue that *MAGEC3* is an X-linked tumor suppressor that escapes X chromosome inactivation.

*MAGEC3* belongs to the melanoma antigen gene (MAGE) family, whose members are defined by a common 200 residue protein domain (protein family 01454) [5] conserved from a single copy in fungi with heavy duplication in mammals and primate-specific clades [6]. Members of the family have varied reports of function in DNA repair, cell cycle control, and protein interaction [7], especially in the context of cancer [8], as well as potential use as stemness markers [9].

Historically, the MAGE family has been studied as *bona fide* tumor specific antigens, and we have previously reported on their individual and combined effects on ovarian cancer prognosis [10], [11]. While tumor expression of most “Type I” tumor specific MAGE genes are associated with poor prognosis, the level of tissue specific expression and clinical implications of MAGEC3 expression are currently unknown. In this work, we measured MAGEC3 protein levels in ovarian cancers, kidney cancers, and normal tissues. Additionally, we measured MAGEC3 levels in samples of various tissue types containing matched RNA sequencing data which were used to build a robust predictor of protein expression. Clinical correlations and molecular associations related to MAGEC3 expression (or loss of expression) elucidate MAGE biology as well as potential therapeutic options.

## RESULTS

### MAGEC3 protein normal tissue expression and cancer tissue expression

In normal tissues, MAGEC3 was widely expressed at a similar level except for decreased expression in breast and skin tissues and increased expression in tonsil, brain, and testis tissues (**Figure 1A**). Normal ovary tissues had a mean expression of 181.2 on a 0-300 point scale (H-score, median 176.5, sd 40.8) while unmatched ovarian cancers had significantly lower expression (mean 145.2, median 135.4, sd 40.4; two-sample t-test, p < 0.001) (**Figure 1B**). Normal kidney tissues had a similar mean expression of 174.9 (sd 20.9) and median expression of 171.6 (**Figure 1D**); non-tumor kidney tissue (not shown) in patients with renal cell carcinoma (RCC) was slightly lower (mean 167.8, median 170.7; two-sample t-test, p=0.003).

**Figure 1.**
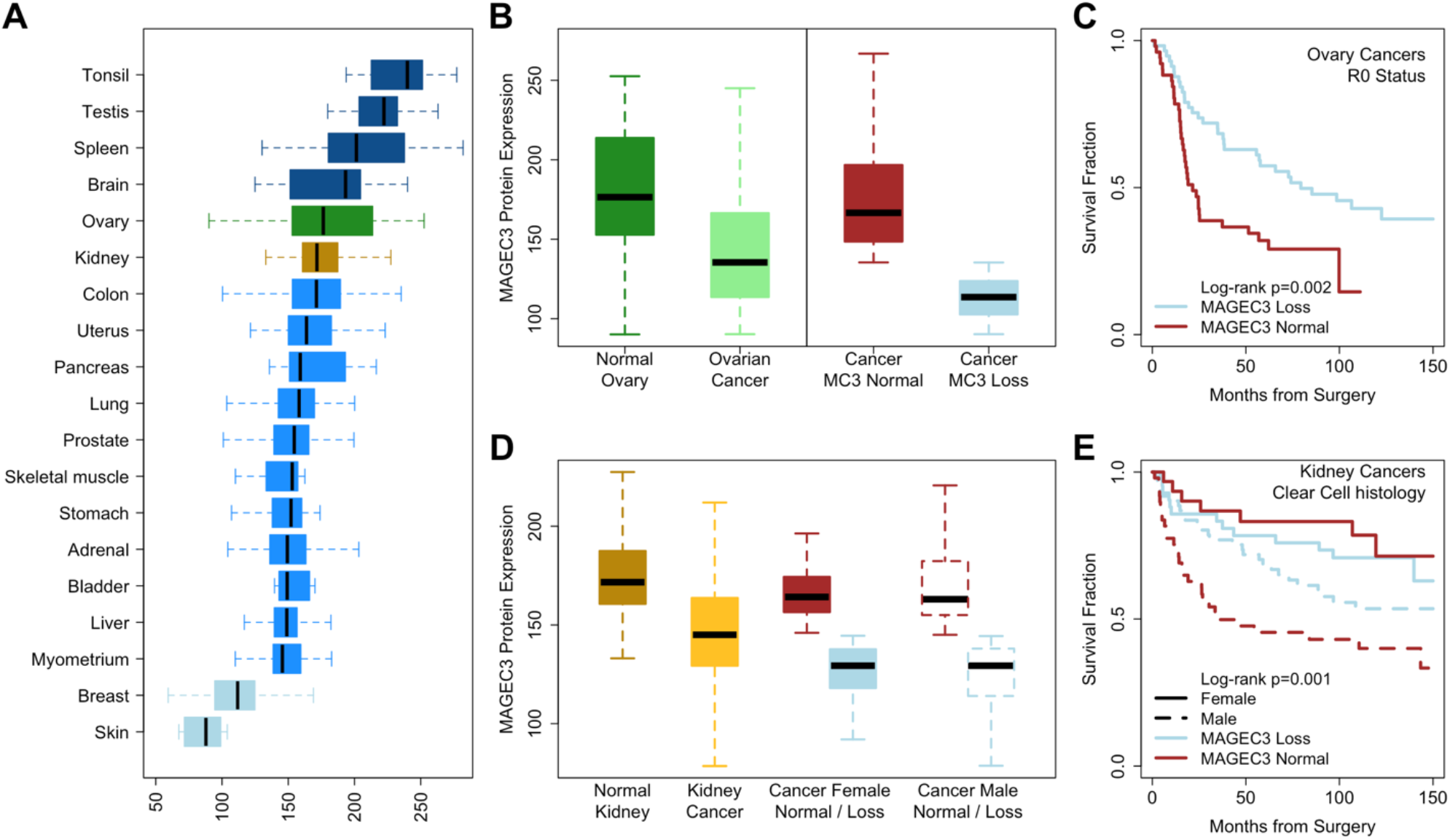
MAGEC3 protein expression in normal and cancer tissues. (A) MAGEC3 protein expression in normal tissue cores. (B) MAGEC3 expression in normal ovary tissue and ovarian cancer (left side). Ovarian cancer expression levels (light green box) were dichotomized at the median into “Normal” (red) and “Loss” (blue) (right side). (C) Kaplan-Meier plot showing progression-free survival trends for ovarian cancers with optimal cytoreduction stratified at the cancer median. (D) MAGEC3 expression in normal kidney tissue and kidney cancer. Kidney cancer expression levels (light yellow box) were dichotomized at the median into “Normal” (red) and “Loss” (blue) and subsequently stratified by sex (filled vs unfilled boxes). (E) Kaplan-Meier plot showing overall survival trends for clear cell kidney cancers stratified by MAGEC3 (median cut) and sex.

Stratified at the median of tumor expression (corresponding to the 9.5th percentile of normal ovary expression), higher levels of MAGEC3 were not different from normal ovary (t-test p=0.368), while lower levels of MAGEC3 were significantly lower (p<0.001), reflecting loss of expression (**Figure 1B**). We subsequently named these tumor groups “Normal” and “Loss”. Normal MAGEC3 levels were associated with cases ascertained after 2006 (OR=5.86, 95%CI: 3.8-9.3, p<0.001) (**Table 1**).

**Table 1.** Clinical characteristics of the ovary discovery cohort by MAGEC3 protein level.

| Characteristic | All Ovary Patients<br>n=411 | MAGEC3 Loss<br>n=206 | MAGEC3 Normal<br>n=205 | P-value |
| --- | --- | --- | --- | --- |
| **Age of Diagnosis [years]** |  |  |  |  |
| Mean (range) | 63 (21-93) | 63 (21-93) | 64 (21-89) | 0.3† |
| Missing | 0 | 0 | 0 |  |
| **Year of Diagnosis [n (%)]** |  |  |  |  |
| Before 2006 | 226 (57.7) | 149 (78.4) | 77 (38.1) | < 0.001‡ |
| After 2006 | 166 (42.3) | 41 (21.6) | 125 (61.9) |  |
| Missing | 19 | 16 | 3 |  |
| **Primary [n (%)]** |  |  |  |  |
| Ovary | 339 (83.5) | 177 (86.8) | 162 (80.2) | 0.1‡ |
| Primary peritoneal | 67 (16.5) | 27 (13.2) | 40 (19.8) |  |
| Missing | 5 | 2 | 3 |  |
| **FIGO Stage [n (%)]** |  |  |  |  |
| I/II/IIIA/B | 70 (17.4) | 41 (20.4) | 29 (14.4) | 0.1‡ |
| IIIC/IV | 333 (82.6) | 160 (79.6) | 173 (85.6) |  |
| Missing | 8 | 5 | 3 |  |
| **Grade [n (%)]** |  |  |  |  |
| Well/Moderately differentiated | 108 (26.7) | 59 (29.4) | 49 (24.1) | 0.3‡ |
| Poorly/Undifferentiated | 296 (73.3) | 142 (70.6) | 154 (75.9) |  |
| Missing | 7 | 5 | 2 |  |
| **Histology [n (%)]** |  |  |  |  |
| Serous | 333 (81.0) | 161 (78.2) | 172 (83.9) | 0.2‡ |
| Other Epithelial | 78 (19.0) | 45 (21.8) | 33 (16.1) |  |
| Missing | 0 | 0 | 0 |  |
| **Cytoreduction [n (%)]** |  |  |  |  |
| R0 | 109 (26.8) | 57 (28.2) | 52 (25.5) | 0.6‡ |
| Not R0 | 297 (73.2) | 145 (71.8) | 152 (74.5) |  |
| Missing | 5 | 4 | 1 |  |
| **Treatment Outcome [n (%)]** |  |  |  |  |
| Complete response | 197 (56.8) | 90 (51.4) | 107 (62.2) | 0.06‡ |
| Not complete response | 150 (43.2) | 85 (48.6) | 65 (37.8) |  |
| Missing | 64 | 31 | 33 |  |
| **Platinum Sensitivity [n (%)]** |  |  |  |  |
| Sensitive | 170 (53.6) | 83 (51.2) | 87 (56.1) | 0.4‡ |
| Resistant | 147 (46.4) | 79 (48.8) | 68 (43.9) |  |
| Missing | 94 | 44 | 50 |  |
| **Survival [months]** |  |  |  |  |
| Median progression-free survival | 18.9 | 24.1 | 16.2 | 0.002§ |
| Median overall survival | 43.0 | 45.8 | 40.2 | 0.2§ |

†P-value was calculated using Student's t-test.
n may vary by characteristic due to missing data
‡P-value was calculated using the chi-squared test.
§P-value was calculated using the log-rank test.

In RCC patients, significantly lower MAGEC3 protein expression was identified in tumors (mean 146.6, median 145.0) compared to matched non-tumor samples (difference mean -22.13, median -23.33, paired t-test, p < 0.001) (**Figure 1D**). MAGEC3 protein levels were associated with black patients (OR=6.70, 95%CI:1.8-47.6, p=0.003) and also with non-clear cell (papillary) histology (OR=14.5, 95%CI: 4.1-99.5, p<0.001) (**Table 2**). Primary tissue expression in cases with synchronous metastases (N=73) was not different from non-metastatic cases (N=147) (148.8 vs 145.7, t-test, p=0.496), and metastatic expression was not different from their matched primaries (N=23, 146.6 vs 146.6, p=0.999).

**Table 2.** Clinical characteristics of the kidney discovery cohort by MAGEC3 protein level.

| Characteristic | All Kidney Patients<br>n=220 | MAGEC3 Loss<br>n=110 | MAGEC3 Normal<br>n=110 | P-value |
| --- | --- | --- | --- | --- |
| **Age at Surgery [years]** |  |  |  |  |
| Mean (range) | 59 (28-89) | 60 (28-87) | 59 (29-89) | 0.4† |
| Missing | 1 | 1 | 0 |  |
| **Sex [n (%)]** |  |  |  |  |
| Male | 136 (61.8) | 67 (60.9) | 69 (62.7) | 0.9‡ |
| Female | 84 (38.2) | 43 (39.1) | 41 (37.3) |  |
| Missing | 0 | 0 | 0 |  |
| **Race [n (%)]** |  |  |  |  |
| Black | 15 (6.8) | 2 (1.8) | 13 (11.8) | 0.008‡ |
| White | 204 (93.2) | 107 (98.2) | 97 (88.2) |  |
| Missing | 1 | 1 | 0 |  |
| **Smoking Status [n (%)]** |  |  |  |  |
| Current | 47 (22.0) | 25 (23.8) | 22 (20.2) | 0.4‡ |
| Former | 69 (32.2) | 29 (27.6) | 40 (36.7) |  |
| Never | 98 (45.8) | 51 (48.6) | 47 (43.1) |  |
| Missing | 6 | 5 | 1 |  |
| **BMI [n (%)]** |  |  |  |  |
| 0 - 24.9 | 36 (18.9) | 13 (13.7) | 23 (24.2) | 0.5‡ |
| 24.9 - 29.9 | 58 (30.5) | 30 (31.6) | 28 (29.5) |  |
| 29.9 - 34.9 | 48 (25.3) | 26 (27.4) | 22 (23.2) |  |
| 34.9 - 39.9 | 30 (15.8) | 16 (16.8) | 14 (14.7) |  |
| > 39.9 | 18 (9.5) | 10 (10.5) | 8 (8.4) |  |
| Missing | 30 | 15 | 15 |  |
| **Histology [n (%)]** |  |  |  |  |
| Clear cell | 184 (83.6) | 104 (94.5) | 80 (72.7) | < 0.001‡ |
| Papillary | 26 (11.8) | 2 (1.8) | 24 (21.8) |  |
| Chromophobe | 10 (4.5) | 4 (3.6) | 6 (5.5) |  |
| Missing | 0 | 0 | 0 |  |
| **Histological Features [n (%)]** |  |  |  |  |
| Clear cell present | 173 (84.4) | 94 (92.2) | 79 (76.7) | 0.004‡ |
| Clear cell absent | 32 (15.6) | 8 (7.8) | 24 (23.3) |  |
| Missing | 15 | 8 | 7 |  |
| Papillary present | 23 (11.2) | 1 (1.0) | 22 (21.4) | < 0.001‡ |
| Papillary absent | 182 (88.8) | 101 (99.0) | 81 (78.6) |  |
| Missing | 15 | 8 | 7 |  |
| Chromophobe present | 9 (4.4) | 3 (2.9) | 6 (5.8) | 0.5‡ |
| Chromophobe absent | 196 (95.6) | 99 (97.1) | 97 (94.2) |  |
| Missing | 15 | 8 | 7 |  |
| Sarcoma present | 17 (8.3) | 7 (6.9) | 10 (9.7) | 0.6‡ |
| Sarcoma absent | 188 (91.7) | 95 (93.1) | 93 (90.3) |  |
| Missing | 15 | 8 | 7 |  |
| **Size [n (%)]** |  |  |  |  |
| 0 – 4 cm | 64 (32.7) | 30 (30.9) | 34 (34.3) | 0.7‡ |
| 4 – 7 cm | 61 (31.1) | 33 (34.0) | 28 (28.3) |  |
| 7 – 10 cm | 42 (21.4) | 22 (22.7) | 20 (20.2) |  |
| > 10 cm | 29 (14.8) | 12 (12.4) | 17 (17.2) |  |
| Missing | 24 | 13 | 11 |  |
| **Metastatic Disease [n (%)]** |  |  |  |  |
| Yes | 73 (33.2) | 36 (32.7) | 37 (33.6) | 1‡ |
| No | 147 (66.8) | 74 (67.3) | 73 (66.4) |  |
| Missing | 0 | 0 | 0 |  |
| **Focality [n (%)]** |  |  |  |  |
| Multifocal | 15 (7.3) | 7 (6.9) | 8 (7.8) | 0.6‡ |
| Unifocal | 148 (72.2) | 71 (69.6) | 77 (74.8) |  |
| Unknown | 42 (20.5) | 24 (23.5) | 18 (17.5) |  |
| Missing | 15 | 8 | 7 |  |
| **Survival [months]** |  |  |  |  |
| Median overall survival | 166.5 | 187.6 | 143.4 | 0.2§ |

†P-value was calculated using Student's t-test.
n may vary by characteristic due to missing data
‡P-value was calculated using the chi-squared test.
§P-value was calculated using the log-rank test.

### MAGEC3 association with prognosis in epithelial ovarian cancer

In ovarian cancers with survival follow up (N=394), normal MAGEC3 levels were associated with complete response (CR) to maximal debulking surgery followed by platinum-based chemotherapy (CR mean 150.5 versus 138.9, two-sample t-test, p=0.008). Following initial complete response, women who were platinum sensitive (more than 9 months of progression-free survival after the end of platinum chemotherapy) continued to show higher MAGEC3 levels (platinum sensitive mean 149.8 versus 140.2, t-test p=0.033). The inverse was seen for progression-free survival (PFS) in all women with epithelial ovarian cancer (normal MAGEC3: 16.2 months median versus 24.1 months, log-rank p=0.002) (**Table 1**). Overall survival (OS) was significantly lower in women with normal MAGEC3 and optimal cytoreduction (R0, 64.7 months versus 118.6, p=0.050) but not in women with suboptimal cytoreduction (not R0, 33.9 versus 34.4, p=0.900).

The age at diagnosis, FIGO stage, grade, and R0 cytoreduction were univariately associated with prognosis in ovarian cancers (**Table 3A**). MAGEC3 level was independently associated with PFS (HR=1.41, 95%CI:1.14-1.75, p=0.002) (**Figure 1C**). Stepwise multivariate model selection by analysis of deviance led to a model that predicted progression-free survival using FIGO Stage (IIIC/IV, HR=3.14, score test p<0.001), histological grade (Poorly/Undifferentiated, HR=1.38, p=0.017), cytoreduction status (not R0, HR=1.45, p=0.022) and MAGEC3 level (Normal, HR=1.41, p=0.004) stratified on histological type and decade of life at diagnosis (global proportional hazards test p=0.32). Women were followed for an average of 52.6 months (maximum 244.4 months). These results were not affected by the year of ascertainment effect (PFS log-rank p=0.40, OS, p=0.20).

**Table 3.**
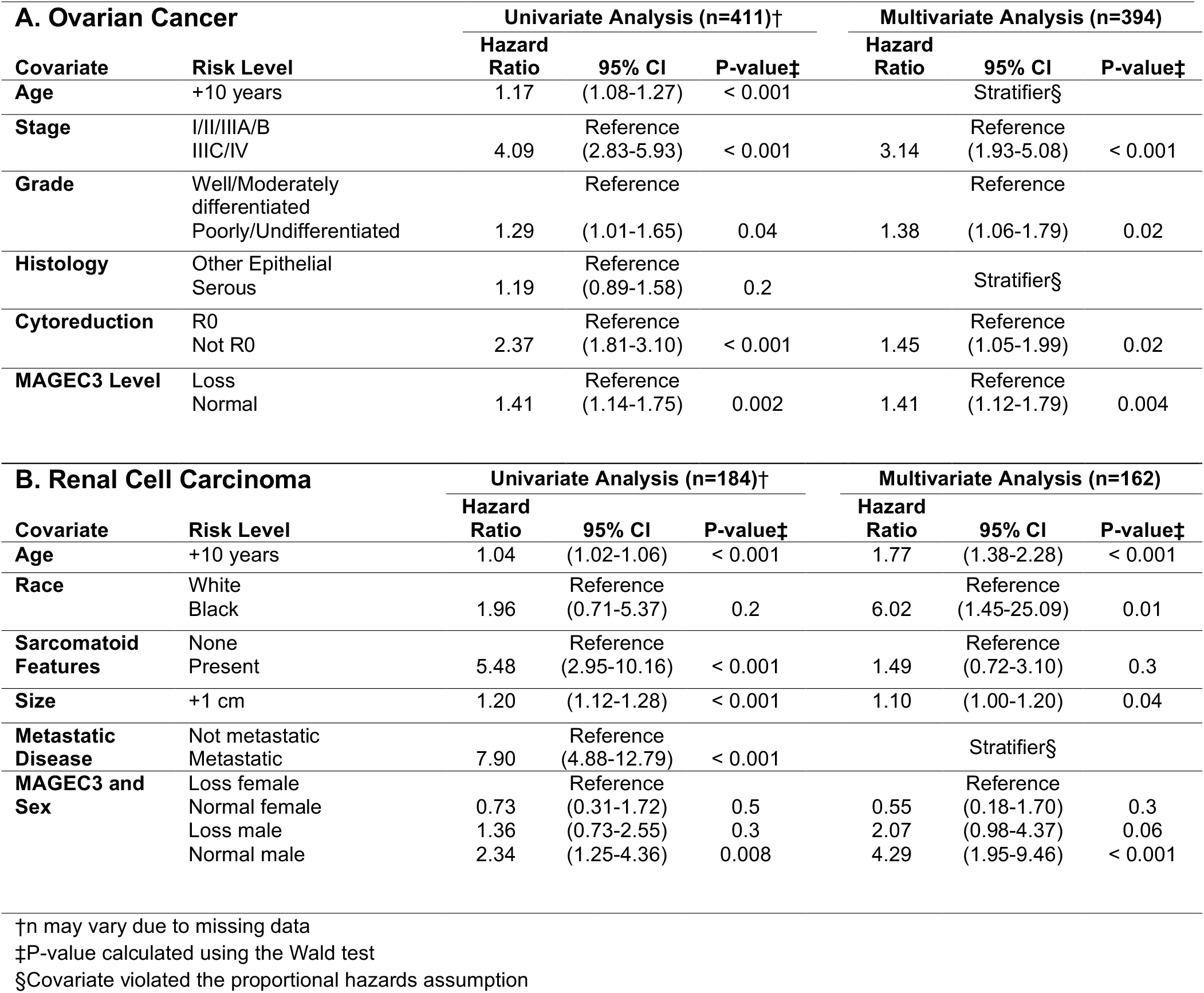
Univariate and multivariate survival analyses of (a) epithelial ovarian cancer (global proportional hazards test p=0.32) and (b) clear cell renal cell carcinoma patients (global proportional hazards test p=0.28).

### MAGEC3 sex-specific effect on overall survival in clear cell renal cell carcinoma

Recalling that MAGEC3 levels were lowest in clear cell tumors among kidney cancers, we focused on only clear cell tumors (N=162) to study a population with expected prognosis similar to that of epithelial ovarian cancers. Among clear cell RCC, overall survival was associated with age at resection, the presence of sarcomatoid features, and synchronous metastases at surgery (**Table 3B**). Additionally, MAGEC3 and sex were jointly associated with survival. With reference to women with MAGEC3 loss, neither women with normal MAGEC3 (HR=0.73, 0.31-1.72, p=0.50) nor men with MAGEC3 loss (HR=1.36, 0.73-2.55, p=0.30) had significantly different hazards. In contrast, men with normal levels had a significantly increased hazard (HR=2.34, 1.25-4.36, p=0.008) (**Figure 1E**).

Stepwise model selection led to a model predicting disease-specific overall survival including, in sequential order (**Table 3B**): histological presence of sarcomatoid cells (HR=1.49, final model p=0.017), age at surgery (+10 years, HR=1.77, p=0.002), size of tumor (+1 cm, HR=1.10, p=0.048), race (black, HR=6.02, p=0.030), and an interaction between sex and normal MAGEC3 level (p<0.001) stratified on the presence of metastasis at surgery (global proportional hazards test p=0.28). The MAGEC3 and sex interaction was interpreted as follows; with a reference level of women with MAGEC3 loss, women with normal MAGEC3 had a non-significant effect (HR=0.55, score test p=0.299), men with MAGEC3 loss were at increased risk (HR=2.07, p=0.056), and men with normal MAGEC3 were at a highly significant increased risk (HR=4.29, p=0.0003).

### MAGEC3 is associated with CD8+ T cell infiltrated tumors

Given that ovarian cancers are highly immunogenic and that kidney tumors are responsive to immune checkpoint therapy, we considered the association between MAGEC3 and the count of CD8+ tumor infiltrating lymphocytes (TILs). We previously reported CD8*+* TIL counts stained in ovarian cancer slides from these TMAs [12]. CD8+ TIL levels were positively correlated with MAGEC3 expression in ovarian tumors (Pearson’s r=0.176, p=0.011) (**Figure S1A**). We similarly stained and quantified the density of CD8+ TILs on the matching kidney TMAs, which had a similar trend (r=0.105, p=0.122) meriting further modeling. Adjusting for histology and the presence of metastatic disease, the correlation coefficient of the correctly scaled linear model was β=0.149 (score test p=0.0344) (**Figure S1B**).

### Relationship with other CT antigens and seropositivity

We have previously reported on the expression of MAGE-A3, MAGE-A4, MAGE-A10, and PRAME [10, 13] in a subset of these ovarian cancers. Among all complete records (N=72), MAGEC3 was not strongly correlated with any MAGE-A antigens (A3, r=0.02, p=0.870; A4, r=0.16, p=0.185; A10, r=-0.06, p=0.604) (**Figure S2A-C**). A larger set of patients (N=195) had complete information on MAGEC3, PRAME (an autosomal antigen) [13] and NY-ESO-1 (gene *CTAG1B* at Xq28) expression and serology [11]. MAGEC3 was strongly correlated with PRAME expression (r=0.422, p<0.001, adjusted for year of diagnosis) (**Figure S2D**) and uncorrelated with NY-ESO-1 expression (r=0.101, p=0.157) (**Figure S2E**). Conversely, patients who were seropositive for NY-ESO-1 had upregulated expression of MAGEC3 (H-score β=11.2, p=0.0323) (**Figure S2F**). Logistic regression predicted NY-ESO-1 seropositivity was specific to MAGEC3 (+10 H-score units, OR=1.26, p=0.035) versus PRAME (OR=1.16, p=0.233).

### MAGEC3 protein expression across multiple disease sites

FFPE blocks were available from cases submitted by Roswell Park to the Cancer Genome Atlas (TCGA) project from several tissue sites (N=180 tumors as a TMA). We stained these cores and analyzed their correlation with bulk mRNA sequencing data. Due to limited sample sizes within tissue sites, we considered a limited correlative analysis of protein expression in this set of patients (**Table S1**). As there is a known association between MAGEC3 and prognosis in ovarian and kidney cancers, we stratified common primary disease sites of the TCGA cohort by MAGEC3 expression. Only colorectal sites had an association with MAGEC3, displaying normal expression more frequently than MAGEC3 loss (p=0.04). When considering the whole cohort, CD8+ cell levels also have a notable association with MAGEC3 expression (CD8+ cell values associated with normal MAGEC3 expression) (p<0.001). Although race was found to be a factor associated with MAGEC3 levels in the kidney discovery cohort, this was not seen across this TCGA cohort (p=0.32).

### MAGEC3 expression is associated with stress-related processes

MAGEC3-associated differentially expressed genes (DEGs, FDR≤0.5) adjusted for tissue site were more frequently upregulated across this TCGA subset (**Figure 2A**). We observed highly upregulated levels for the rest of the MAGEC locus (*MAGEC1*, and *MAGEC2* on Xq27.2) as well as the neighboring MAGEA locus (*MAGEA2, MAGEA3, MAGEA6, MAGEA10*, and *MAGEA12* on Xq28). Among these MAGE proteins, *MAGEA2* and *MAGEC2* interact with *TRIM28* affecting the cell cycle among other processes [14, 15]. MAGEC3 also upregulates a stress related gene, *WRNIP1* [16]; a DNA replication gene, *MCM7* [17]; and cancer related genes, *XPO5* [18] and *ZSCAN16* [19]. In addition, MAGEC3 downregulates genes that are shown to induce tumor progression including *APLMR* [20, 21], *FBLN5* [22], and *FNDC1* [23, 24]. Gene set enrichment analysis (GSEA) showed that MAGEC3 significantly enriched E2F targets (NES=3.63, FDR<0.001), G2/M checkpoint (NES=3.20, FDR<0.001), and DNA repair (NES=2.28, FDR<0.001), whereas the epithelial to mesenchymal transition was downregulated (NES=-3.34, FDR<0.001) (**Figure 2B**). We also observed that differentially expressed genes were clustered in specific genomic locations such as 11p15, 11p23, 12p13, 17q21, and Xq28 (**Figure 2C**). These data suggest that MAGEC3 may induce a transcriptional response that involves the expression of stress-related genes that affect cell cycle and DNA repair, while downregulating genes involved in tumor progression.

**Figure 2.**
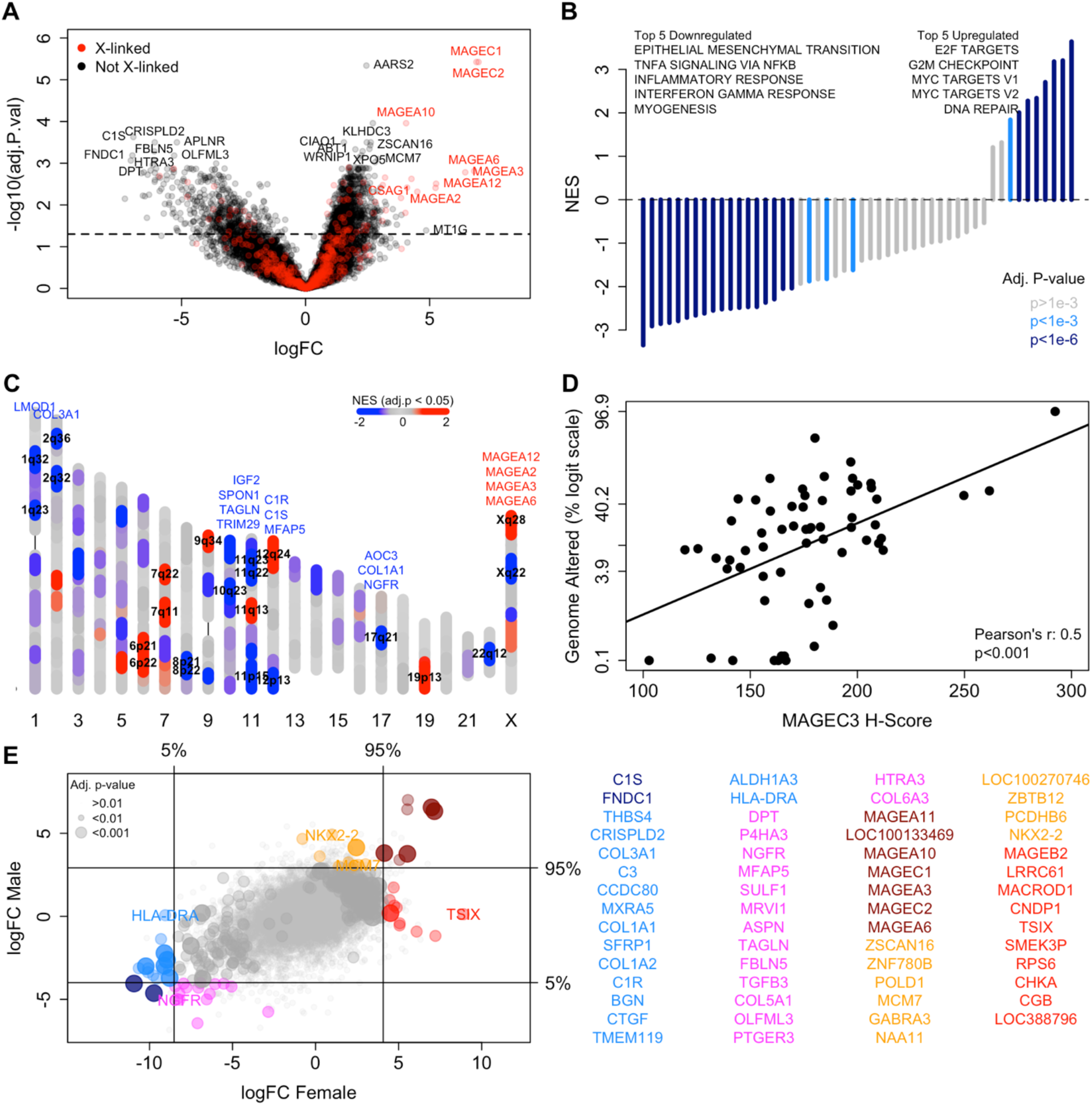
MAGEC3 protein DEG and GSEA analyses and association with genomic instability. (A) Volcano plot showing genes that are differentially expressed with continuous MAGEC3 protein expression. (B) Gene sets that are significantly associated with continuous MAGEC3 protein expression. (C) Ideogram showing where significantly expressed genes are located within the genome. Genes that were individually significant are listed above their locus and loci that are GSEA significant (FDR ≤ 0.05) are labelled. (D) Scatterplot showing the correlation between fraction genome altered and MAGEC3 protein levels in patients with *PTEN, FAT4, BRAF, PTPRT, NF1, RB1, ATM, ATRX, IDH1*, and *TP63* mutations. (E) Comparison of DEGs between male and female patients.

### MAGEC3 expression is correlated with genomic instability in patients with mutated oncogenes

Due to MAGEC3’s association with specific expression patterns in various genomic locations and upregulation of DNA repair processes, we investigated the association of MAGEC3 with genomic instability. We analyzed tumor mutation burden (TMB) data from 63 patients within the TCGA patient cohort stratified on the most mutated genes. A positive correlation between levels of MAGEC3 protein and fraction genome altered (Pearson’s r = 0.5, p<0.001) was observed in patients with *PTEN, FAT4, BRAF, PTPRT, NF1, RB1, ATM, ATRX, IDH1*, and *TP63* mutations (**Figure 2D**). This association was negligible in patients without these oncogenic mutations (Pearson’s r = 0.03, p<0.001) (**Figure S3**).

### MAGEC3 expression associates with different genes depending on sex

Stratifying by sex, we determined which DEGs were male or female specific or common to both groups (**Figure 2E**). Of particular interest was *TSIX*, a long non-coding RNA that inhibits *XIST* expression, thereby preventing X chromosome inactivation [25]. We observed a large log-fold change of *TSIX* expression in female patients with MAGEC3 protein expression indicative of a link between MAGEC3 protein expression and general X-chromosome inactivation. Another female specific DEG is the MHC class II gene *HLA-DRA*, which was specifically downregulated [26]. Male-specific DEGs include *MCM7*, a DNA replication licensing factor [17]; *NKX2-2*, an important biomarker for metastatic prostate cancers [27]; and *NGFR*, the canonical single-pass transmembrane receptor linked to melanoma [28] and squamous cell carcinoma [29].

### Modeling MAGEC3 protein via mRNA

Much like other MAGE genes and X-linked cancer testis antigens, *MAGEC3* had unusually low RNA counts despite measurable protein expression and demonstrated some linear correlation with protein expression (Pearson’s r=0.267, p=0.0003) likely due to a few high leverage points (Spearman’s ρ=-0.003, p=0.968). This observation suggested that, while some high expressing cases could be detected, RNA sequencing lacks the sensitivity to detect *MAGEC3* mRNA levels.

To maximize the utility of mRNA datasets, we modeled protein expression of MAGEC3 using other mRNA measurements and divided the N=180 labeled TCGA set (**Table S1**) into training and validation datasets (2:1 ratio) where the TCGA clear cell kidney (KIRC) [30] and ovary (OV) [31] datasets were considered an unlabeled validation cohort. The final model selected by the 1 standard error (se) rule had a training mean square error (MSE) of 455.5 (**Figure 3A**) and a leave out one test MSE of 637.9 (N=120, se: 110.11). The fully withheld validation set MSE was 885.3 (N=60). Correlation between true MAGEC3 protein scores and predicted scores was 0.69 (p<0.001) in the model building cohort and 0.51 (p<0.001) in the validation cohort (**Figure 3B-C**). Similar error rates and correlations, such as those identified within these cohorts, imply a well-fit model with predictive modeling capabilities. The fitted model selected mRNA expression of *AMBN, CCDC77, RNF175, C7orf51, MCART3P, CBLL1*, and *KIAA1429* as positively correlated with MAGEC3 protein and *RPS6KA2, LYVE1, CCDC80, CX3CR1, EMP1, CDSN*, and *ANGPT4* as negatively correlated with MAGEC3 protein.

**Figure 3.**
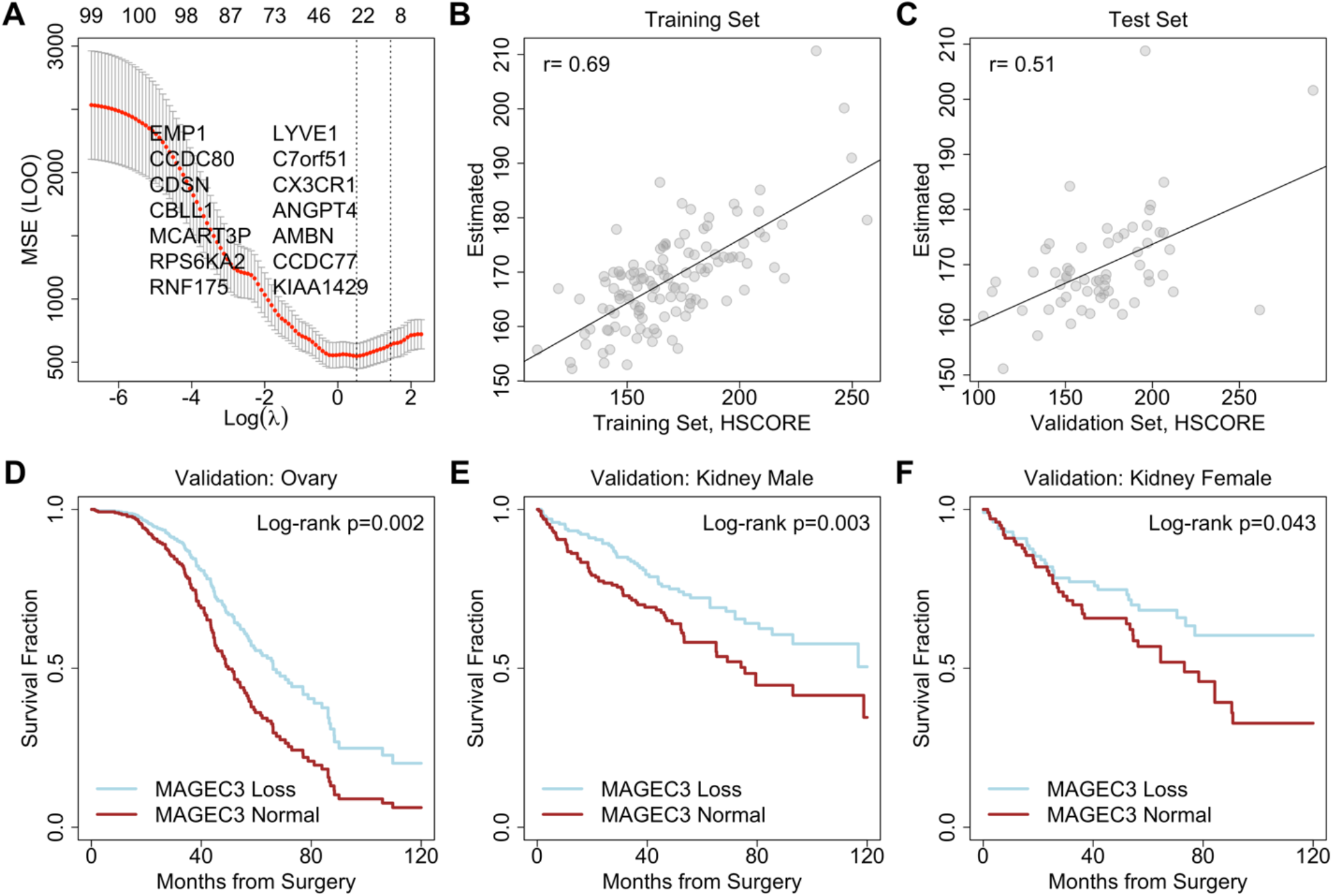
RNA-based predictive model for MAGEC3 expression. (A) Leave out one (LOO) cross-validation error for tuning parameter selection. Fitted values in the (B) training set and (C) withheld test set. Survival estimate validation in the independent pan-TCGA dataset for (D) ovarian cancers, (E) kidney cancer males, and (F) kidney cancer females.

### MAGEC3 protein predictor in Pan-TCGA study

We applied the MAGEC3 protein level predictor to an independent set of cases sequenced and processed for TCGA’s pan-cancer analysis (N=282 ovary, N=606 clear cell RCC) which did not have measured MAGEC3 protein levels. Analyses performed on our discovery cohorts were also performed on these predicted MAGEC3 scores. In ovarian cancers, progression-free survival had a significant negative relationship with normal MAGEC3 scores (HR=1.35 p=0.048) which persisted after accounting for age, stage, and initial response to chemotherapy (HR =1.74 p=0.002). When split at the median, higher MAGEC3 cases had a median survival of 36.9 months (34.8-43.7) versus 51.3 months (95% interval: 45.0-62.1) (**Figure 3D**). In clear cell RCCs, men with high MAGEC3 levels had significantly poorer overall survival (log-rank p=0.003) (**Figure 3E**). Under proportional hazards, the effect (HR=1.67, p=0.004) was of similar magnitude in women (HR=1.62, p=0.045) (**Figure 3F**).

## DISCUSSION

To our knowledge, this is the first investigation of the primate specific MAGEC3 protein expression and its relation to cancer. MAGEC3 appears to be expressed in multiple tissues, including the tonsil, spleen, brain, and testes at a similar level. Normal ovary and kidney tissues had relatively high expression, while MAGEC3 levels were unusually low in breast and skin tissues. This makes MAGEC3 similar to Type II MAGE family members with ubiquitous expression and to the updated classification, “Type 1c: not restricted” [32].

Consistent with the supposition that it is a tumor suppressor, MAGEC3 levels were lower on average in ovarian cancers than in normal ovary tissue and also decreased significantly in matched renal cell carcinomas. Other reports studying *MAGEC3* mRNA (using Affymetrix oligonucleotide arrays, probe 216592_at) show associations with positive prognosis in hepatocellular carcinoma [33] and mixed relapse free survival based on lymph node status in breast cancer [34].

While more work is required to assess sensitivity, specificity, and prevalence, higher relative MAGEC3 protein was a significant biomarker for poor prognosis in both ovary and kidney cancers. Specifically, cases with loss of MAGEC3 expression had unusually good prognosis. We interpret this as evidence that MAGEC3-driven disease etiology is driven by loss of MAGEC3’s protective expression. The favorable prognosis of these cases is consistent with the observation that women with germline *BRCA1* and *BRCA2* loss of function mutations have a favorable [35] but transient [36] response to platinum-based chemotherapy. In contrast, those cases with higher MAGEC3 expression and poor prognosis likely reflect a MAGE-independent form of disease (and not a pro-tumor function for MAGEC3) because the level of tumor expression is not significantly increased above normal tissue.

We observed that specific genes associated with MAGEC3 protein expression were clustered around G2/M checkpoint and DNA repair hallmark pathways, which is consistent with reports that other MAGE family members impact the cell cycle (*MAGEC2*) [37], damage sensing via *p53* targeting E3-ligases (*MAGEC2*) [38] or ATM/ATR phosphorylation (*MAGED2*) [39], and genome stability (*MAGEG1*) [40], all of which are associated with stress response [41]. An interesting conjecture emerges that these non-MAGE driven, high MAGEC3 expressing tumors may retain their MAGE-related putative DNA repair functions. In addition, differentially expressed genes associated with MAGEC3 were clustered in specific genome locations indicating that MAGEC3 may affect chromatin accessibility due to a stress related program [42] that can be related to cell cycle and DNA repair.

Tumor mutational burden is often correlated with proteins associated with DNA damage repair [43]. MAGEC3 protein levels were positively associated with fraction genome altered in patients with *PTEN, FAT4, BRAF, PTPRT, NF1, RB1, ATM, ATRX, IDH1*, and *TP63* mutations. Because this strong correlation with fraction genome altered was only observed in patients with these driver mutations, we believe that expression of MAGEC3 is secondary to the mutations. The tumors may be expressing MAGEC3 in response to high genomic instability caused by their driver mutations.

We reported that *MAGEC3* was difficult to measure by bulk RNA sequencing and subsequently developed an RNA-based linear model to predict MAGEC3 protein levels learned from labelled data. The accuracy of the predictor was evaluated in a validation set and subsequently applied to ovarian cancer and clear cell RCC patients within the TCGA pan-cancer dataset. Using the predicted scores, we were able to validate the results found for cases with directly measured MAGEC3 scoring. The ability to accurately predict protein levels based off an RNA predictor greatly increased the sample size and impact of our results.

MAGE proteins are cancer-testis antigens [44]. MAGEC3 protein was associated with increased expression of mRNA in the MAGE-C locus (at Xq27) and the MAGE-A locus (at Xq28). Interestingly, MAGEC3 was a good predictor for seropositive ovarian cancer patients even though NY-ESO-1 (at Xq28) mRNA and protein were not associated with MAGEC3 expression. These results, along with the correlation of MAGEC3 protein and CD8 positivity, indicate that MAGEC3 may be related to the re-opening of Xq28 and the expression of tumor antigens.

A limitation of our investigation involves the use of retrospective cohorts. We did observe a tissue processing effect for FFPE blocks obtained prior to 2006 that did not significantly affect our conclusions after stratification and sensitivity analysis. The use of immunohistochemistry to read the MAGE protein levels is semi-quantitative and slide-to-slide variation was minimized using normal controls on each tissue microarray and automatic staining in an experienced core pathology facility. This protocol was used in previous work with other stains. The MAGEC3 antibody was vetted by the Human Protein Atlas [45] and proven to be appropriate for IHC.

Despite these limitations, our study used a large single-institution cohort of patients with definitive power to provide a first investigation of MAGEC3 protein expression in ovarian and kidney cancer cases and to confirm the gene’s relevance to cancer biology and patient prognosis. In an independent, model-building cohort and a further validation cohort, MAGEC3 was identified as a candidate prognostic biomarker associated with both ovarian and kidney cancers with the potential for predictive marker-based treatment strategies based on targeting cases with high expression.

## METHODS

### Patient cohorts and TMA construction

Three cohorts of patients were used in this study: (1) a “discovery cohort” of ovarian and kidney cancer patients from Roswell Park, (2) a “model-building” cohort of mutually exclusive patients from Roswell Park that had been accrued and molecularly characterized through the TCGA project and (3) a second validation cohort of TCGA patients from other institutions without protein data.

With respect to the first cohort, following review by the Roswell Park Institutional Review Board, we obtained clinical data and archival samples for: (a) women with a primary diagnosis of ovarian cancer treated by maximal debulking surgery and first line platinum-based chemotherapy and (b) patients with a primary diagnosis of renal cell carcinoma undergoing nephrectomy at the institution. We required that patients in both cohorts had available tumor blocks taken prior to the initiation of systemic therapy. In total, there were N=220 RCC patients and N=411 ovarian cancer patients (high-grade serous ovarian cancer, N=333) for analysis. Tissue blocks were organized into tissue microarrays as previously described [10, 46] and stained for MAGEC3 protein expression. TMAs contained reference cores for normalization across different blocks.

### Antibody validation

The MAGEC3 antibody was vetted by the Human Protein Atlas [45] (HPA052067) following the guidelines of the International Working Group for Antibody Validation [47] and was scored as approved for IHC and supported for antigen specificity. The antigen appears to bind both dominant isoforms of *MAGEC3* (ESNP000000386566 and ENSP00000440444) outside of the functional MAGE homology domain.

### Immunohistochemistry

Formalin-fixed paraffin sections were cut at 4µm, placed on charged slides, and dried at 60°C for one hour. Slides were cooled to room temperature and added to the Dako Omnis autostainer, where they were deparaffinized with Clearify (American Mastertech; catalog# CACLEGAL) and rinsed in water. Flex TRS High pH (Dako; catalog# GV804) was used for target retrieval for 60 minutes. Slides were incubated with MAGEC3 antibody (Sigma Aldrich; catalog# HPA052067) for 50 minutes at 1:50 dilution. Flex Rabbit Linker (Dako; catalog# GV80911-2) was applied for 10 minutes followed by Flex /HRP polymer (Dako; catalog# DM843) which was applied for 20 minutes followed by DAB (Diaminobenzidine) (Dako; catalog# K3468) for 5 minutes for visualization. Slides were counterstained with Hematoxylin for 8 minutes then put into water. After removing slides from the Omnis they are dehydrated, cleared, and cover slipped. Cores were averaged by patient and checked for outliers and slides were normalized to common internal control spots included in the design of the TMA. MAGEC3 was quantified using an IHC Profiler [48]. CD8 (Dako #M7103) was stained as previously described [12]. MAGEA3, MAGEA4 and MAGEA10 were stained as previously described [10] and scored as positive or negative. PRAME was stained and quantitatively scored as previously described [13]. After staining, slide-specific staining intensity and rank and file associations were tested to identify any slide/batch or spatial effects.

### TCGA study data

We downloaded clinical and expression data for the TCGA Pan-Cancer (PANCAN) cohort directly from UCSC Xena browser as batch effect normalized mRNA data (log2 RSEM, upper-quartile normalized [49]). Modifications made to the data after download include the removal of unknown or hypothetical transcripts in the Entrez database and the setting of NA counts to 0. Corresponding clinical information for the expression data were obtained from PANCAN’s curated clinical data [50].

### Statistical analysis

Statistical tests are described as they are used in text. In general, Student’s t-test, chi-square test and log-rank test were employed as appropriate for continuous, categorical or survival time data. All statistical tests were two-sided and p<0.05 was considered significant throughout. Survival analysis by Cox’s proportional hazards model used analysis of deviance-based model selection and employed residual tests for non-proportionality [51]. Overall survival was defined as a cancer associated death event censored by end of follow up. For ovarian cancers, progression-free survival was defined as doubling of baseline CA125 confirmed with CT scan or the initiation of relapse chemotherapy regimens as defined in [52]. All analyses were performed in the R statistical programming language version 3.6.1.

### LASSO model building

Within the model-building cohort, genes with consistent univariate association were screened for MAGEC3 association and the top 1000 features with univariate association with MAGEC3 [53] were selected for multivariate model building. We used a LASSO-penalized linear model objective to select and estimate coefficients and selected the tuning parameter by unbiased leave-out-one (LOO) cross validation via R/glmnet [54]. Suboptimal local solutions were ruled out by multiple randomized partitions of the model/validation cohorts.

DEG calling was performed in R/limma using the moderated t-statistics and FDR control [55]. The linear model included the TCGA study as a covariate. Gene set analysis was performed using the R/fgsea package [56]. Annotations were maintained via R/org.Hs.eg.db and MSigDb’s Hallmark pathway set. Pathways with fewer than 15 represented genes were removed from consideration.

## Data Availability

Protein scoring and clinical data for the discovery cohort is available upon request. Clinical and expression data for the TCGA Pan-Cancer (PANCAN) cohort was downloaded from UCSC Xena browser.

## Conflicts of Interest

The authors declare that they have no conflicts of interest.

## Acknowledgements

The results here are in whole or part based upon data generated by the TCGA Research Network: https://www.cancer.gov/tcga. This work was funded by Department of Defense grants OC170368 and PC180449 (to KHE), the Roswell Park Cancer Center Support Grant P30CA016056, and grants from the Roswell Park Alliance Foundation.

## SUPPLEMENTAL TABLES AND FIGURES

**Supplemental Table 1.** Clinical characteristics of TCGA validation cohort by MAGEC3 protein level.

| Characteristic | All Patients<br>n=180 | MAGEC3 Loss<br>n=90 | MAGEC3 Normal<br>n=90 | P-value |
| --- | --- | --- | --- | --- |
| **Age of Diagnosis [years]** |  |  |  |  |
| Mean (range) | 60 (24-90) | 60 (24-90) | 60 (30-90) | 0.97† |
| Missing | 1 | 1 | 0 |  |
| **Year of Diagnosis [n (%)]** |  |  |  |  |
| Before 2006 | 49 (27.4) | 29 (32.6) | 20 (22.2) | 0.17‡ |
| After 2006 | 130 (72.6) | 60 (67.4) | 70 (77.8) |  |
| Missing | 1 | 1 | 0 |  |
| **Primary Site\* [n (%)]** |  |  |  |  |
| Colorectal | 28 (23.0) | 10 (16.7) | 18 (29.0) | 0.04‡ |
| Kidney | 17 (13.9) | 11 (18.3) | 6 (9.7) | 0.23‡ |
| Breast | 14 (11.5) | 11 (18.3) | 3 (4.8) | 0.10‡ |
| Lung | 14 (11.5) | 9 (15.0) | 5 (8.1) | 0.61‡ |
| Sarcomas | 7 (5.7) | 1 (1.7) | 6 (9.7) | 0.82‡ |
| Prostate | 6 (4.9) | 5 (8.3) | 1 (1.6) | 0.30‡ |
| Ovary | 5 (4.1) | 1 (1.7) | 4 (6.5) | 0.27‡ |
| Melanoma | 5 (4.1) | 2 (3.3) | 3 (4.8) | 1.00‡ |
| Thyroid | 5 (4.1) | 4 (6.7) | 1 (1.6) | 0.47‡ |
| Other | 21 (17.2) | 6 (10.0) | 15 (24.2) |  |
| Missing | 58 | 30 | 28 |  |
| **Sex [n (%)]** |  |  |  |  |
| Male | 102 (57.0) | 48 (53.9) | 54 (60.0) | 0.50‡ |
| Female | 77 (43.0) | 41 (46.1) | 36 (40.0) |  |
| Missing | 1 | 1 | 0 |  |
| **Race [n (%)]** |  |  |  |  |
| Black or African American | 23 (13.8) | 14 (17.1) | 9 (10.6) | 0.32‡ |
| White | 142 (85.0) | 67 (81.7) | 75 (88.2) |  |
| Other | 2 (1.2) | 1 (1.2) | 1 (1.2) |  |
| Missing | 13 | 8 | 5 |  |
| **AJCC Tumor Stage [n (%)]** |  |  |  |  |
| I | 28 (29.5) | 18 (35.3) | 10 (22.7) | 0.66‡ |
| II | 25 (26.3) | 12 (23.5) | 13 (29.6) |  |
| III | 27 (28.4) | 10 (19.6) | 17 (38.6) |  |
| IV | 15 (15.8) | 11 (21.6) | 4 (9.1) |  |
| Missing | 85 | 39 | 46 |  |
| **CD8 Level [% positive area]** |  |  |  |  |
| Mean (range) | 36 (11-77) | 30 (11-60) | 42 (22-77) | < 0.001† |
| Missing | 34 | 17 | 17 |  |
| **Survival [months]** |  |  |  |  |
| Median progression-free survival | 72.0 | 84.3 | 71.5 | 0.62§ |
| Median overall survival | 117.9 | NA | 117.9 | 0.51§ |

†P-value was calculated using Student's t-test.
‡P-value was calculated using the chi-squared test.
§P-value was calculated using the log-rank test.
NA-median not achieved
\*rare sites: lymph node, uterus, endometrium, urinary bladder, adrenal gland, liver, omentum, abdominal wall, cervix, mesentery, myometrium, pancreas, spleen, stomach, thymus
n may vary by characteristic due to missing data

**Supplemental Figure 1.**
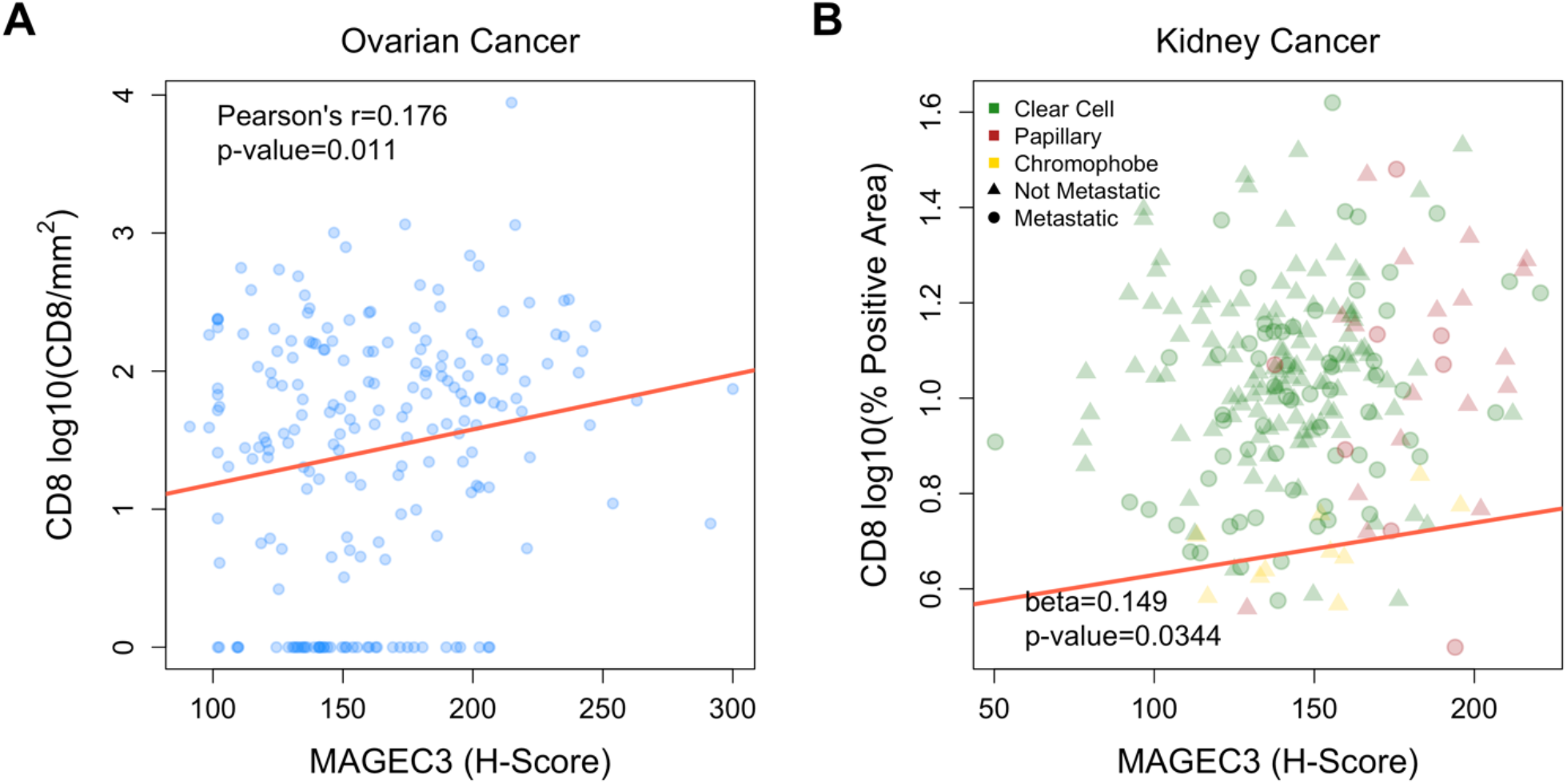
TMAs were stained and scored to determine protein levels of MAGEC3 (H-score) and CD8 (stain intensity per mm^2^ and percent positive area). Plotting those levels reveals a positive correlation between MAGEC3 and CD8 in (A) ovarian cancer cases and also (B) kidney cancer cases after adjustment for histology and presence of metastatic disease.

**Supplemental Figure 2.**
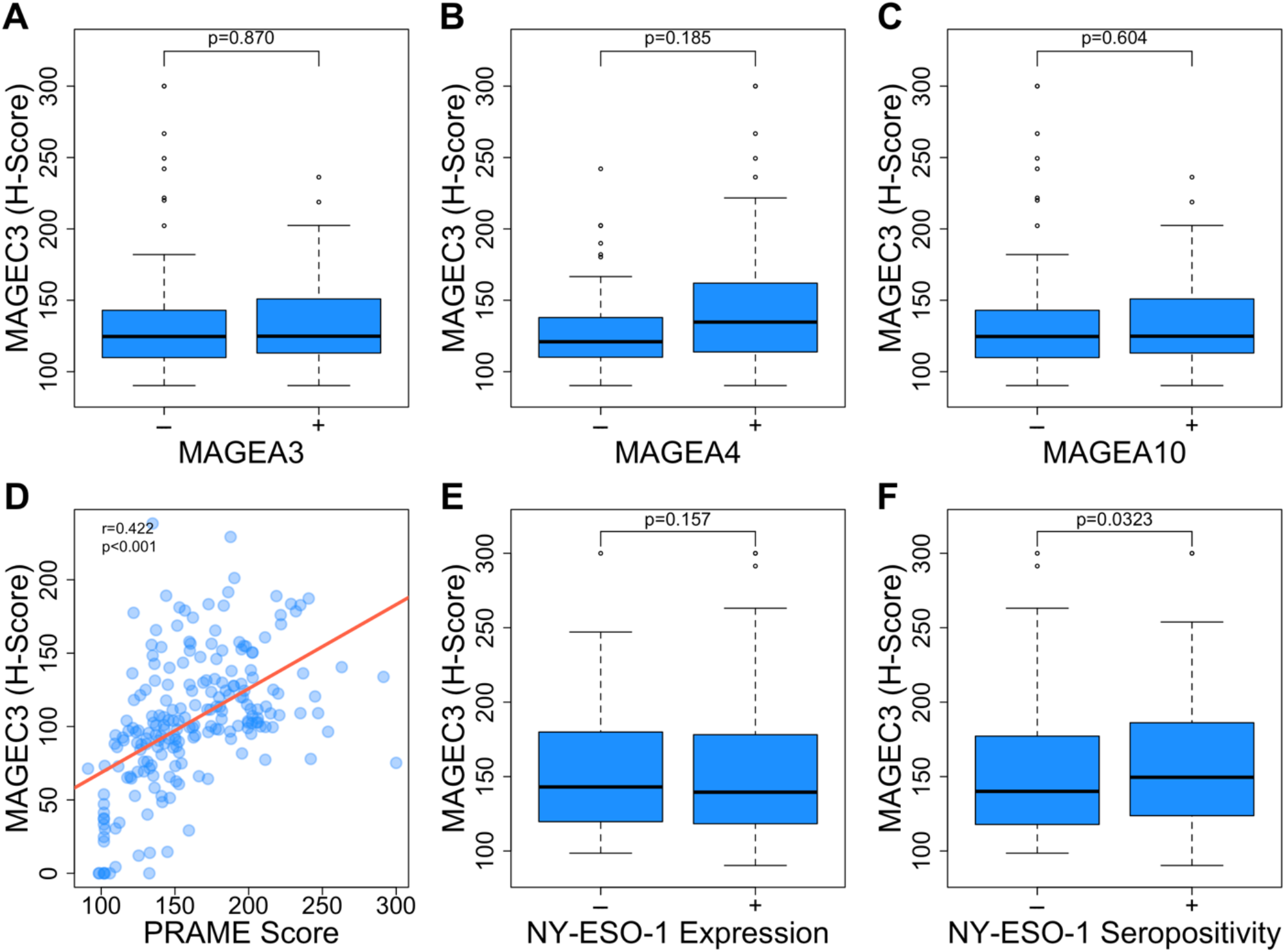
A subset of 72 ovarian cancer patients within our discovery cohort contained binary expression data for MAGE-A antigens. There is no significant correlation between MAGEC3 and (A) MAGEA3, (B) MAGEA4, or (C) MAGEA10. A slightly larger set of 195 ovarian cancer patients had data for PRAME expression, NY-ESO-1 expression, and NY-ESO-1 seropositivity. There is a positive correlation between MAGEC3 and (D) PRAME levels as well as (F) NY-ESO-1 seropositivity, but not between MAGEC3 and (E) NY-ESO-1 expression.

**Supplemental Figure 3.**
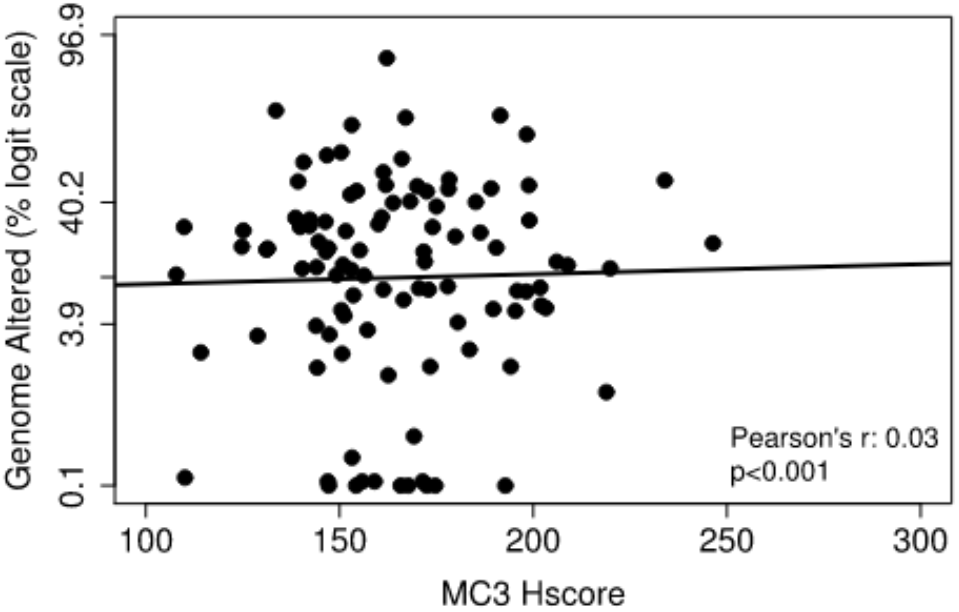
Scatterplot showing that fraction genome altered is not correlated with MAGEC3 protein levels in patients lacking *PTEN, FAT4, BRAF, PTPRT, NF1, RB1, ATM, ATRX, IDH1*, or *TP63* mutations.

## Notes

### Competing Interest Statement

The authors have declared no competing interest.

### Author Declarations

The use of clinical data and archival samples was reviewed and approved by the Roswell Park Institutional Review Board.

## REFERENCES

1. Eng, K.H., et al., Paternal lineage early onset hereditary ovarian cancers: A Familial Ovarian Cancer Registry study. PLoS Genet, 2018. 14(2): p. e1007194.

2. Piver, M.S., Hereditary ovarian cancer. Lessons from the first twenty years of the Gilda Radner Familial Ovarian Cancer Registry. Gynecol Oncol, 2002. 85(1): p. 9–17.

3. Etter, J.L., et al., Transmission of X-linked Ovarian Cancer: Characterization and Implications. Diagnostics (Basel), 2020. 10(2).

4. Dunford, A., et al., Tumor-suppressor genes that escape from X-inactivation contribute to cancer sex bias. Nat Genet, 2017. 49(1): p. 10–16.

5. Chomez, P., et al., An overview of the MAGE gene family with the identification of all human members of the family. Cancer Res, 2001. 61(14): p. 5544–51.

6. Katsura, Y. and Y. Satta, Evolutionary history of the cancer immunity antigen MAGE gene family. PLoS One, 2011. 6(6): p. e20365.

7. Florke Gee, R.R., et al., Emerging Roles of the MAGE Protein Family in Stress Response Pathways. J Biol Chem, 2020.

8. Weon, J.L. and P.R. Potts, The MAGE protein family and cancer. Curr Opin Cell Biol, 2015. 37: p. 1–8.

9. Gordeeva, O., A. Gordeev, and S. Khaydukov, Expression dynamics of Mage family genes during self-renewal and differentiation of mouse pluripotent stem and teratocarcinoma cells. Oncotarget, 2019. 10(35): p. 3248–3266.

10. Daudi, S., et al., Expression and immune responses to MAGE antigens predict survival in epithelial ovarian cancer. PLoS One, 2014. 9(8): p. e104099.

11. Szender, J.B., et al., NY-ESO-1 expression predicts an aggressive phenotype of ovarian cancer. Gynecol Oncol, 2017. 145(3): p. 420–425.

12. Tsuji, T., et al., Clonality and antigen-specific responses shape the prognostic effects of tumorinfiltrating T cells in ovarian cancer. Oncotarget, 2020. 11(27): p. 2669–2683.

13. Zhang, W., et al., PRAME expression and promoter hypomethylation in epithelial ovarian cancer. Oncotarget, 2016. 7(29): p. 45352–45369.

14. Marcar, L., et al., MAGE-A Cancer/Testis Antigens Inhibit MDM2 Ubiquitylation Function and Promote Increased Levels of MDM4. PLoS One, 2015. 10(5): p. e0127713.

15. Doyle, J.M., et al., MAGE-RING protein complexes comprise a family of E3 ubiquitin ligases. Mol Cell, 2010. 39(6): p. 963–74.

16. Socha, A., et al., WRNIP1 Is Recruited to DNA Interstrand Crosslinks and Promotes Repair. Cell Rep, 2020. 32(1): p. 107850.

17. Qu, K., et al., MCM7 promotes cancer progression through cyclin D1-dependent signaling and serves as a prognostic marker for patients with hepatocellular carcinoma. Cell Death Dis, 2017. 8(2): p. e2603.

18. Lin, D., et al., Exportin-5 SUMOylation promotes hepatocellular carcinoma progression. Exp Cell Res, 2020. 395(2): p. 112219.

19. Li, X., et al., ZSCAN16 promotes proliferation, migration and invasion of bladder cancer via regulating NF-kB, AKT, mTOR, P38 and other genes. Biomed Pharmacother, 2020. 126: p. 110066.

20. PodgOrska, M., et al., The Role of Apelin and Apelin Receptor Expression in Migration and Invasiveness of Colon Cancer Cells. Anticancer Res, 2021. 41(1): p. 151–161.

21. Masoumi, J., et al., Role of Apelin/APJ axis in cancer development and progression. Adv Med Sci, 2020. 65(1): p. 202–213.

22. Yuan, R., et al., LOXL1 exerts oncogenesis and stimulates angiogenesis through the LOXL1-FBLN5/alphavbeta3 integrin/FAK-MAPK axis in ICC. Mol Ther Nucleic Acids, 2021. 23: p. 797–810.

23. Jiang, T., et al., FNDC1 Promotes the Invasiveness of Gastric Cancer via Wnt/beta-Catenin Signaling Pathway and Correlates With Peritoneal Metastasis and Prognosis. Front Oncol, 2020. 10: p. 590492.

24. Liu, Y.P., et al., Overexpression of FNDC1 Relates to Poor Prognosis and Its Knockdown Impairs Cell Invasion and Migration in Gastric Cancer. Technol Cancer Res Treat, 2019. 18: p. 1533033819869928.

25. Gayen, S., et al., A Primary Role for the Tsix lncRNA in Maintaining Random X-Chromosome Inactivation. Cell Rep, 2015. 11(8): p. 1251–65.

26. Rangel, L.B., et al., Anomalous expression of the HLA-DR alpha and beta chains in ovarian and other cancers. Cancer Biol Ther, 2004. 3(10): p. 1021–7.

27. Xue, M., et al., Computational identification of mutually exclusive transcriptional drivers dysregulating metastatic microRNAs in prostate cancer. Nat Commun, 2017. 8: p. 14917.

28. Boshuizen, J., et al., Reversal of pre-existing NGFR-driven tumor and immune therapy resistance. Nat Commun, 2020. 11(1): p. 3946.

29. Elkashty, O.A., et al., Cancer stem cells enrichment with surface markers CD271 and CD44 in human head and neck squamous cell carcinomas. Carcinogenesis, 2020. 41(4): p. 458–466.

30. Cancer Genome Atlas Research, N., et al., Comprehensive Molecular Characterization of Papillary Renal-Cell Carcinoma. N Engl J Med, 2016. 374(2): p. 135–45.

31. Berger, A.C., et al., A Comprehensive Pan-Cancer Molecular Study of Gynecologic and Breast Cancers. Cancer Cell, 2018. 33(4): p. 690–705 e9.

32. Fon Tacer, K., et al., MAGE cancer-testis antigens protect the mammalian germline under environmental stress. Sci Adv, 2019. 5(5): p. eaav4832.

33. Li, R., et al., A comprehensive analysis of the MAGE family as prognostic and diagnostic markers for hepatocellular carcinoma. Genomics, 2020.

34. Jia, B., et al., Prognostic roles of MAGE family members in breast cancer based on KM-Plotter Data. Oncol Lett, 2019. 18(4): p. 3501–3516.

35. Zhong, Q., et al., Effects of BRCA1-and BRCA2-related mutations on ovarian and breast cancer survival: a meta-analysis. Clin Cancer Res, 2015. 21(1): p. 211–20.

36. Kotsopoulos, J., et al., Ten-year survival after epithelial ovarian cancer is not associated with BRCA mutation status. Gynecol Oncol, 2016. 140(1): p. 42–7.

37. Hao, J., et al., Cancer-testis antigen MAGE-C2 binds Rbx1 and inhibits ubiquitin ligase-mediated turnover of cyclin E. Oncotarget, 2015. 6(39): p. 42028–39.

38. Xiao, T.Z., et al., MAGE I transcription factors regulate KAP1 and KRAB domain zinc finger transcription factor mediated gene repression. PLoS One, 2011. 6(8): p. e23747.

39. Pirlot, C., et al., Melanoma antigen-D2: A nucleolar protein undergoing delocalization during cell cycle and after cellular stress. Biochim Biophys Acta, 2016. 1863(4): p. 581–95.

40. Taylor, E.M., et al., Identification of the proteins, including MAGEG1, that make up the human SMC5-6 protein complex. Mol Cell Biol, 2008. 28(4): p. 1197–206.

41. Florke Gee, R.R., et al., Emerging roles of the MAGE protein family in stress response pathways. J Biol Chem, 2020. 295(47): p. 16121–16155.

42. Seddon, A.R., et al., Genome-wide impact of hydrogen peroxide on maintenance DNA methylation in replicating cells. Epigenetics Chromatin, 2021. 14(1): p. 17.

43. Birkbak, N.J., et al., Tumor mutation burden forecasts outcome in ovarian cancer with BRCA1 or BRCA2 mutations. PLoS One, 2013. 8(11): p. e80023.

44. Li, X.F., et al., The expression, modulation and use of cancer-testis antigens as potential biomarkers for cancer immunotherapy. Am J Transl Res, 2020. 12(11): p. 7002–7019.

45. Uhlen, M., et al., A human protein atlas for normal and cancer tissues based on antibody proteomics. Mol Cell Proteomics, 2005. 4(12): p. 1920–32.

46. Greene, C.J., et al., Transferrin receptor 1 upregulation in primary tumor and downregulation in benign kidney is associated with progression and mortality in renal cell carcinoma patients. Oncotarget, 2017. 8(63): p. 107052–107075.

47. Uhlen, M., et al., A proposal for validation of antibodies. Nat Methods, 2016. 13(10): p. 823–7.

48. Varghese, F., et al., IHC Profiler: an open source plugin for the quantitative evaluation and automated scoring of immunohistochemistry images of human tissue samples. PLoS One, 2014. 9(5): p. e96801.

49. Bullard, J.H., et al., Evaluation of statistical methods for normalization and differential expression in mRNA-Seq experiments. BMC Bioinformatics, 2010. 11: p. 94.

50. Liu, J., et al., An Integrated TCGA Pan-Cancer Clinical Data Resource to Drive High-Quality Survival Outcome Analytics. Cell, 2018. 173(2): p. 400–416 e11.

51. Grambsch, P.T. T., Proportional hazards tests and diagnostics based on weighted residuals. Biometrika, 1994. 81: p. 515–526.

52. Starbuck, K.D., et al., Prognostic impact of adjuvant chemotherapy treatment intensity for ovarian cancer. PLoS One, 2018. 13(11): p. e0206913.

53. Fan, J., R. Samworth, and Y. Wu, Ultrahigh dimensional feature selection: beyond the linear model. J Mach Learn Res, 2009. 10: p. 2013–2038.

54. Friedman, J.H. T; Tibshirani, R, Regularization Paths for Generalized Linear Models via Coordinate Descent. Journal of Statistical Software, 2010. 33(1): p. 1–22.

55. Ritchie, M.E., et al., limma powers differential expression analyses for RNA-sequencing and microarray studies. Nucleic Acids Res, 2015. 43(7): p. e47.

56. Sergushichev, A., An algorithm for fast preranked gene set enrichment analysis using cumulative statistic calculation. BbioRxiv, 2016.

